# High-sensitivity C-reactive protein profiles differ between Korean and US adults with LDL cholesterol below 100 mg/dL

**DOI:** 10.64898/2026.10.14.26365531

**Authors:** Jun Teng, Junyan Xia

**Affiliations:** Department of Traditional Chinese Medicine, The Second Affiliated Hospital, Zhejiang University School of Medicine, 88 Jiefang Road, Shangcheng District, Hangzhou 310009, Zhejiang, China; Department of Traditional Chinese Medicine, Sir Run Run Shaw Hospital, Zhejiang University School of Medicine, 3 East Qingchun Road, Shangcheng District, Hangzhou 310016, Zhejiang, China

**Keywords:** High-sensitivity C-reactive protein, Low-density lipoprotein cholesterol, Adiposity, Population surveillance, Health examination surveys

## Abstract

**Objectives:** To compare high-sensitivity C-reactive protein (hsCRP) profiles in Korean and US adults with low-density lipoprotein cholesterol (LDL-C) below 100 mg/dL and examine differences after adiposity adjustment.

**Study design:** Cross-sectional analysis of national health examination surveys.

**Methods:** We analysed the Korea National Health and Nutrition Examination Survey 2022–2024 (n=5,785) and US National Health and Nutrition Examination Survey August 2021–August 2023 (n=1,056). Eligible adults had Sampson-calculated LDL-C <100 mg/dL and hsCRP ≤10 mg/L; elevated hsCRP was defined as 2–10 mg/L. We estimated survey-weighted prevalence differences and adjusted prevalence ratios (PRs).

**Results:** LDL-C distributions closely overlapped (weighted medians, 79.7 vs 80.1 mg/dL), whereas median hsCRP was 0.50 vs 1.34 mg/L. Elevated-hsCRP prevalence was 12.2% in Korea and 35.7% in the United States (difference, 23.5 percentage points; 95% CI, 19.4–27.7); the age–sex-standardised difference was 23.4 percentage points. Among 6,235 common complete cases, the age–sex-adjusted US-versus-Korea PR was 2.86 (95% CI, 2.48–3.30), decreasing to 1.66 (1.40–1.98) after waist circumference adjustment and 1.92 (1.62–2.26) after body mass index adjustment.

**Conclusions:** Similar LDL-C distributions coexisted with markedly different hsCRP profiles. Waist adjustment attenuated the between-country contrast more than BMI adjustment, supporting joint consideration of inflammatory, lipid and anthropometric measures in population surveillance.

## 1. Introduction

Although lowering low-density lipoprotein cholesterol (LDL-C) remains central to cardiovascular prevention, lipid measurements do not capture systemic inflammatory activity. The 2026 ACC/AHA dyslipidaemia guideline identifies LDL-C <100 mg/dL as a goal for selected primary-prevention adults and persistently elevated high-sensitivity C-reactive protein (hsCRP) as a risk enhancer.^1^ In a pooled analysis of 31,245 statin-treated trial participants, hsCRP was more strongly associated with subsequent cardiovascular events and mortality than LDL-C.^2^ Yet the frequency of elevated hsCRP within this LDL-C range, and its variation across national populations, remain less well characterised.

National health examination surveys offer a broader perspective than clinical treatment cohorts. An analysis of the 2024 Korea National Health and Nutrition Examination Survey (KNHANES) reported hsCRP ≥2 mg/L in 11.7% of adults without conventional lipid abnormalities.^3^ This finding illustrates discordance between lipid classification and inflammatory status, but conventional lipid-normal definitions encompass LDL-C concentrations above 100 mg/dL. Furthermore, findings from one national population cannot establish whether this discordance is similarly frequent elsewhere. Whether populations with comparable LDL-C distributions also have comparable hsCRP profiles therefore remains uncertain. Resolving this question would inform interpretation of cardiometabolic population surveillance beyond conventional lipid measures alone.

Adiposity provides a plausible context for such variation. Adipose-tissue dysfunction is associated with systemic inflammation,^4^ whereas body mass index (BMI) cannot adequately characterise fat distribution.^5^ Waist circumference provides complementary information about abdominal adiposity, potentially distinguishing populations with similar overall body size but different inflammatory profiles. These distinctions are relevant when comparing countries whose anthropometric characteristics may differ even within a common LDL-C range. We therefore compared hsCRP distributions and elevated-hsCRP prevalence in Korean and US adults with LDL-C <100 mg/dL and examined how demographic characteristics and measures of overall and abdominal adiposity relate to the between-country contrast.

## 2. Methods

We conducted a cross-sectional analysis of KNHANES 2022–2024 and the US National Health and Nutrition Examination Survey (NHANES) August 2021–August 2023, both stratified, multistage probability surveys. Eligible adults were aged ≥20 years, had positive analysis weights and valid survey-design information, had fasted for 8 to <24 hours, and had valid lipid and hsCRP measurements. LDL-C was calculated in both surveys from total cholesterol, high-density lipoprotein cholesterol (HDL-C) and triglycerides using the Sampson equation.^6^ Triglycerides were restricted to >0 and <800 mg/dL; LDL-C eligibility was based on unrounded values. The primary sample comprised adults with calculated LDL-C <100 mg/dL and hsCRP ≤10 mg/L. Elevated hsCRP was defined as 2–10 mg/L, with lower values retained in the denominator (Supplementary Figure S1).

Principal covariates were age and sex, with waist circumference and body mass index (BMI) analysed as continuous measures of adiposity. Additional cardiometabolic covariates included triglycerides, HDL-C, glucose, glycated haemoglobin, systolic blood pressure, estimated glomerular filtration rate, diabetes and hypertension. Lifestyle/treatment variables comprised current smoking, current alcohol use and a lipid-lowering treatment proxy. Survey-specific variable definitions and measurement procedures are provided in the Supplementary Methods and Table S1. Released hsCRP values were analysed without cross-survey assay calibration.

### 2.1 Statistical analysis

Within each survey, estimates incorporated sampling weights, strata and primary sampling units. KNHANES examination weights were divided by three for the combined survey years; NHANES used fasting subsample weights (WTSAF2YR). We summarised weighted participant characteristics, cumulative LDL-C and hsCRP distributions, and elevated-hsCRP prevalence. Continuous characteristics were reported as weighted means (SDs) or medians (IQRs), and categorical characteristics as weighted percentages. Variances were estimated by Taylor linearisation; US-minus-Korea prevalence differences used summed independent-survey variances and normal-approximation 95% confidence intervals (CIs).

Direct age–sex standardisation used four age groups (20–39, 40–59, 60–69 and ≥70 years) crossed with sex. The reference distribution was the average of the two surveys’ weighted age–sex proportions, treated as fixed for variance estimation. US-versus-Korea prevalence ratios (PRs) were estimated using survey-weighted log-link quasi-Poisson regression with design-based CIs.^7^ Regression weights were normalised to a mean of one within each survey, with distinct survey/year stratum and cluster identifiers.

All models were fitted to a common complete-case sample defined by the full covariate set, including BMI, to allow comparisons without changes in sample composition. Following unadjusted and age–sex-adjusted models, separate models added waist circumference or BMI, followed by a joint adiposity model. The waist model was then extended sequentially with cardiometabolic and lifestyle/treatment variables. Continuous covariates were standardised, with triglycerides log-transformed first. Missing covariates were not imputed. Sensitivity analyses varied lipid definitions, the hsCRP upper cutoff, survey-period alignment, treatment-proxy restriction, complete-case selection and relative survey weighting (Supplementary Tables S2, S4 and S5). Analyses used R 4.6.0; further specifications appear in the Supplementary Methods.

## 3. Results

The primary sample comprised 5,785 Korean and 1,056 US adults, including 724 and 394 with elevated hsCRP, respectively (Supplementary Figure S1). US participants were younger (mean age, 49.2 vs 52.9 years) and had higher BMI (28.6 vs 24.4 kg/m^2^) and waist circumference (98.5 vs 85.1 cm), whereas lipid-lowering treatment-proxy prevalence was similar (37.6% vs 36.9%; Supplementary Table S3). Weighted median LDL-C was nearly identical in Korea and the United States (79.7 vs 80.1 mg/dL), with closely overlapping cumulative distributions. In contrast, median hsCRP was 0.50 mg/L (IQR, 0.30–1.00) in Korea and 1.34 mg/L (IQR, 0.58–2.96) in the United States (Figure 1A–B).

**Figure 1.**
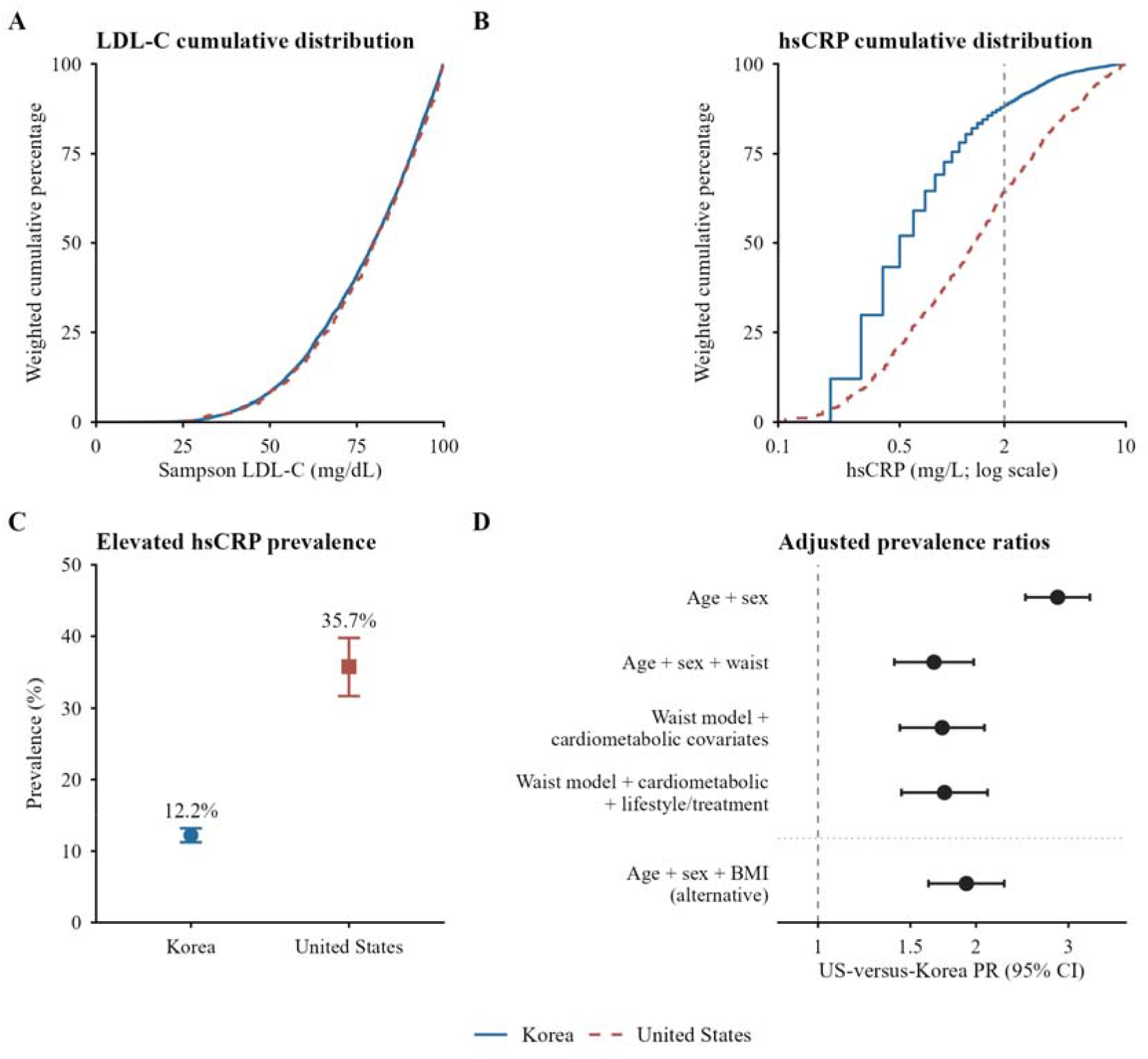
Lipid and inflammatory profiles and adjusted prevalence ratios in Korea and the United States. **A**, Survey-weighted cumulative Sampson LDL-C distribution. **B**, Survey-weighted cumulative hsCRP distribution on a logarithmic axis; the grey dashed line marks 2 mg/L. **C**, Elevated-hsCRP prevalence with 95% confidence intervals (CIs); Korea is shown by a blue circle and the United States by a brick-red square. Curves use blue solid and brick-red dashed lines, respectively. A–C use the primary samples (Korea, n=5,785; United States, n=1,056). **D**, US-versus-Korea prevalence ratios (PRs) and design-based 95% CIs in common complete cases (N=6,235; 972 with elevated hsCRP). Cardiometabolic and lifestyle/treatment adjustments extend the waist model; BMI is an alternative to waist. Elevated hsCRP is 2–10 mg/L among adults with Sampson LDL-C <100 mg/dL and hsCRP ≤10 mg/L. BMI, body mass index; hsCRP, high-sensitivity C-reactive protein; LDL-C, low-density lipoprotein cholesterol.

Weighted elevated-hsCRP prevalence was 12.2% (95% CI, 11.2–13.2) in Korea and 35.7% (31.7–39.8) in the United States, an absolute difference of 23.5 percentage points (19.4–27.7; Figure 1C). Age–sex standardisation left the difference essentially unchanged (12.6% vs 36.0%; difference, 23.4 percentage points [19.4–27.5]; Supplementary Table S4).

Among 6,235 common complete cases (972 with elevated hsCRP), the age–sex-adjusted US-versus-Korea prevalence ratio (PR) was 2.86 (95% CI, 2.48–3.30). Adjustment for waist circumference reduced the PR to 1.66 (1.40–1.98), compared with 1.92 (1.62–2.26) after BMI adjustment. Joint waist and BMI adjustment yielded 1.68 (1.41–2.01). The waist-based PR remained similar after additional cardiometabolic adjustment (1.72 [1.43–2.07]) and subsequent lifestyle/treatment adjustment (1.74 [1.44–2.10]; Figure 1D; Supplementary Table S5).

The absolute difference persisted across alternative lipid criteria, including survey-provided LDL-C <100 mg/dL, calculated LDL-C <70 mg/dL and non-HDL-C <130 mg/dL (range, 22.4–24.5 percentage points). Without the hsCRP upper cutoff, the difference was 27.1 percentage points (95% CI, 22.6–31.6); restricting Korean data to 2022–2023 yielded 24.0 (19.8–28.2), and restricting analysis to adults without the treatment proxy yielded 20.1 (13.9–26.3). Model-specific complete cases and alternative survey weighting produced contrasts in the same direction (Supplementary Tables S4 and S5).

## 4. Discussion

In these national health examination surveys, Korean and US adults within the same LDL-C range had closely overlapping LDL-C distributions but markedly different hsCRP profiles. Elevated hsCRP was 23.5 percentage points more prevalent in the United States, and age–sex standardisation changed this contrast little. Separation across the cumulative hsCRP distributions showed that the finding was not confined to a single threshold. Adiposity adjustment narrowed the between-country contrast, but a considerable difference persisted after broader adjustment.

These findings extend evidence that lipid and inflammatory measures capture distinct dimensions of cardiometabolic health. In statin-treated clinical trials, hsCRP carried prognostic information independently of LDL-C,^2^ and a recent Korean survey identified elevated hsCRP among adults without conventional lipid abnormalities.^3^ Our cross-national comparison adds that similar LDL-C distributions can coexist with substantially different inflammatory profiles. The prevalence contrast persisted under alternative lipid definitions, including non-HDL-C, and among participants without the lipid-lowering treatment proxy. This is relevant because lipid-lowering therapies may also affect CRP concentrations.^8^

Adiposity provided an additional perspective. The age–sex-adjusted prevalence ratio declined from 2.86 to 1.66 after waist adjustment, compared with 1.92 after BMI adjustment; joint adjustment yielded 1.68. Waist circumference reflects abdominal size, which BMI does not capture,^5^ while adipose-tissue dysfunction and immune activation are associated with systemic inflammation.^4^ An earlier analysis of KNHANES 2022 likewise reported attenuation of the hsCRP–metabolic syndrome association after waist adjustment.^9^ This finding provides additional context for the different adjustment patterns observed for waist circumference and BMI. The persisting country contrast after further cardiometabolic and lifestyle/treatment adjustment also suggests that the measured variables do not fully characterise the difference.

The public health implication is that relatively low LDL-C concentrations need not correspond to low prevalence of inflammatory biomarker elevation. Joint surveillance of lipids and hsCRP can distinguish the frequency above a shared threshold from shifts across the full biomarker distribution. Reporting waist circumference alongside BMI also provides a more informative description of differences in population adiposity. These measures are already collected in both national surveys, making such integrated reporting feasible without introducing new population examinations.

National probability sampling, a shared LDL-C equation and multiple sensitivity analyses strengthened the comparison. Important limitations include the absence of cross-survey hsCRP assay calibration, different anatomical sites for waist measurement and the lack of fasting-specific KNHANES weights comparable to NHANES weights. Single hsCRP measurements are subject to within-person variability,^10^ and the treatment proxy does not fully characterise medication exposure. Future studies using repeated national surveys and harmonised biomarker and anthropometric measurements could establish whether these differences persist over time and clarify the relationship with abdominal adiposity. Together, these findings support considering inflammatory and anthropometric profiles alongside lipids in population cardiometabolic surveillance.

## Ethical approval

KNHANES and NHANES obtained ethics approval and participant informed consent under their respective survey protocols. The present study was a secondary analysis of publicly available, deidentified survey data and involved no direct participant contact or intervention.

## Acknowledgements

The authors thank the participants and survey teams of KNHANES and NHANES for their contributions and acknowledge the respective agencies for making the survey data publicly available.

## Author contributions

Jun Teng: Conceptualization, Methodology, Software, Formal analysis, Visualization, Writing–original draft. Junyan Xia: Data curation, Validation, Writing–original draft, Writing–review and editing. Both authors read and approved the final manuscript.

## Data availability

KNHANES 2022–2024 public-use data are available after registration from the Korea Disease Control and Prevention Agency (https://knhanes.kdca.go.kr/knhanes/main.do). NHANES August 2021–August 2023 data are available from the US Centers for Disease Control and Prevention (https://wwwn.cdc.gov/nchs/nhanes/continuousnhanes/default.aspx?Cycle=2021-2023).

## Declaration of generative AI and AI-assisted technologies in the manuscript preparation process

During the preparation of this work, the authors used ChatGPT(OpenAI) to improve language and readability. After using these tools, the authors reviewed and edited the content as needed and take full responsibility for the content of the published article.

