## Supplementary Material for "High-sensitivity C-reactive protein profiles differ between Korean and US adults with LDL cholesterol below 100 mg/dL"

**Supplementary Methods**

**A. Data preparation and variable definitions**

Records were linked by SEQN in NHANES and participant ID plus year in KNHANES. Shared eligibility and hsCRP definitions are specified in the main Methods. Sampson inputs required total cholesterol (TC)>0, high-density lipoprotein cholesterol (HDL-C)>0 and 0<triglycerides (TG)<800 mg/dL, with TC≥HDL-C. LDL-C = TC/0.948 − HDL-C/0.971 − (TG/8.56 + TG×(TC−HDL-C)/2140 − TG²/16100) − 9.44, with lipid inputs in mg/dL (main reference 6; supplementary reference S1). Classification used unrounded values.

Released hsCRP values were used without cross-survey calibration. Assays, lower limits, instrument changes and waist measurement sites are listed in Table S1; Korean 2022–2023 analytical lower limits remain unconfirmed. Diabetes combined reported diagnosis/treatment, fasting glucose ≥126 mg/dL or glycated haemoglobin (HbA1c) ≥6.5%; hypertension combined diagnosis/treatment, systolic pressure ≥130 mmHg or diastolic pressure ≥80 mmHg. Estimated glomerular filtration rate (eGFR) used the 2021 race-free CKD-EPI creatinine equation (supplementary reference S4). Survey-specific medication, smoking and alcohol variables are summarized in Table S1. The medication proxy does not identify drug class, dose or adherence.

**B. Survey analysis**

NHANES used WTSAF2YR, SDMVSTRA and SDMVPSU; KNHANES used WT_ITVEX/3, KSTRATA and PSU. Strata combined survey, year and source stratum; cluster identifiers additionally included source PSU. R survey::svydesign used nest=TRUE, with survey.lonely.psu="adjust". KNHANES examination weights have no fasting-specific counterpart. Variances used Taylor linearization. Independent-survey prevalence differences used summed variances and normal 95% CIs.

Age-sex standardization used the average of the two surveys’ weighted cell proportions; reference weights were fixed for variance estimation. Pooled regression weights had mean 1 within each survey and analyzed sample. PRs were estimated using survey::svyglm with quasipoisson(link="log") and design-based CIs (main reference 7). Cardiometabolic and lifestyle/treatment models extended the waist model. Continuous covariates entered linearly, with triglycerides log-transformed.

**C. Missing data and sensitivity analyses**

Common complete cases required all M0–M4 covariates, including BMI for the alternative and joint-adiposity models, and valid design/weight information (Table S2). Model-specific cases required only the corresponding model’s covariates. No missing-value imputation was performed.

Sensitivity analyses used survey-provided LDL-C <100 mg/dL, Sampson LDL-C <70 mg/dL, non-HDL-C <130 mg/dL, hsCRP ≥2 mg/L without an upper cutoff, Korean 2022–2023 data, or treatment-proxy=0 (Table S4). Shared fasting and valid-lipid restrictions were retained. Equal-total survey weights were evaluated in model-specific age-sex cases (Table S5).

**Table S1. Measurement and variable comparability**

| **Domain** | **KNHANES** | **NHANES** |
| --- | --- | --- |
| LDL-C | Common Sampson calculation: HE_chol, HE_HDL_st2, HE_TG. Native: direct HE_LDL_drct. | Common Sampson calculation: LBXTC, LBDHDD, LBXTLG. Native: Friedewald LBDLDL. |
| hsCRP | HE_hs_crp (mg/L). 2024: particle-enhanced immunoturbidimetry, Cobas 8000; reportable lower limit 0.2 mg/L. Earlier-year analytical limits unconfirmed. | LBXHSCRP (mg/L). Latex-enhanced immunoturbidimetry; Cobas 6000→8000 transition. LOD 0.15 mg/L; below-LOD released value 0.15/√2. |
| Waist / BMI | HE_wc (cm): midpoint between lowest rib and iliac crest. HE_BMI (kg/m²). | BMXWAIST (cm): upper iliac-crest site. BMXBMI (kg/m²). |
| Fasting | HE_fst: 8 to <24 hours; examination weights. | PHAFSTHR: 8 to <24 hours; morning fasting-subsample weights. |
| Survey design | WT_ITVEX/3; KSTRATA; PSU. Years 2022–2024. | WTSAF2YR; SDMVSTRA; SDMVPSU. August 2021–August 2023. |
| Diagnosis / treatment | Diabetes: DE1_dg, DE1_pt, DE1_31. Hypertension: DI1_dg, DI1_pt. Medication proxy: DI2_dg, DI2_pt, DI2_2; code-defined response rules. | Diabetes: DIQ010, DIQ050, DIQ070. Hypertension: BPQ020, BPQ150. Medication proxy: BPQ101D=1→1; =2→0; otherwise missing. |
| Current smoking | BS3_1=1/2→1; =0/3/4/8→0. | SMQ020=2→0; SMQ020=1 with SMQ040=1/2→1 or SMQ040=3→0. |
| Current alcohol use | BD1_11=2–6→1; =1/8→0. | ALQ111=2 or ALQ121=0→0; otherwise ALQ121=1–10→1. |

Smoking and alcohol codes follow the frozen analysis; other responses are missing. Clinical thresholds are specified in the Supplementary Methods. TC, total cholesterol; TG, triglycerides; HDL-C, high-density lipoprotein cholesterol; LOD, limit of detection. Supplementary references S1–S3 and main reference 3. The Korean 0.2 mg/L value is a 2024 reportable limit, not an assumed analytical LOD for all years.

**Table S2. Covariate missingness and complete-case retention**

| **Variable** | **Korea (N=5,785)** | **United States (N=1,056)** |
| --- | --- | --- |
| BMI | 132 (2.3%) | 15 (1.4%) |
| Waist circumference | 191 (3.3%) | 51 (4.8%) |
| Glycated haemoglobin | 12 (0.2%) | 0 (0.0%) |
| Systolic blood pressure | 85 (1.5%) | 45 (4.3%) |
| eGFR | 0 (0.0%) | 9 (0.9%) |
| Hypertension | 65 (1.1%) | 25 (2.4%) |
| Current smoking | 106 (1.8%) | 1 (0.1%) |
| Current alcohol use | 99 (1.7%) | 143 (13.5%) |
| Lipid-lowering treatment proxy | 1 (0.0%) | 9 (0.9%) |

Unweighted missing n (%). Age, sex, triglycerides, HDL-C, glucose and diabetes had no missing values in either survey. Of 6,841 primary participants, 6,235 common complete cases included 972 elevated-hsCRP observations; 606 participants were excluded for missing covariates, including 146 with elevated hsCRP. Variable-specific missing counts overlap.

**Table S3. Participant characteristics**

| **Characteristic** | **Korea (n=5,785)** | **United States (n=1,056)** |
| --- | --- | --- |
| Age, years | 52.9 (17.9) | 49.2 (19.7) |
| Female sex | 3,201 (50.4%) | 539 (48.4%) |
| BMI, kg/m² | 24.4 (3.8) | 28.6 (7.1) |
| Waist circumference, cm | 85.1 (11.2) | 98.5 (17.4) |
| Sampson LDL-C, mg/dL | 79.7 (64.9–90.8) | 80.1 (66.2–91.4) |
| hsCRP, mg/L | 0.50 (0.30–1.00) | 1.34 (0.58–2.96) |
| Triglycerides, mg/dL | 100.0 (72.0–146.0) | 85.0 (59.0–124.0) |
| HDL-C, mg/dL | 56.7 (15.5) | 53.7 (15.0) |
| Non-HDL-C, mg/dL | 98.9 (22.4) | 96.6 (20.4) |
| eGFR, mL/min/1.73 m² | 98.2 (18.3) | 95.8 (23.3) |
| Diabetes | 1,522 (23.2%) | 296 (22.5%) |
| Hypertension | 3,411 (54.3%) | 618 (50.1%) |
| Lipid-lowering treatment proxy | 2,436 (36.9%) | 512 (37.6%) |

Values are survey-weighted mean (SD), except LDL-C, hsCRP and triglycerides, which are weighted median (IQR); categorical values are unweighted n (weighted %). Variable-specific available observations were used. BMI, body mass index; eGFR, estimated glomerular filtration rate; HDL-C, high-density lipoprotein cholesterol; LDL-C, low-density lipoprotein cholesterol; hsCRP, high-sensitivity C-reactive protein; SD, standard deviation; IQR, interquartile range.

**Table S4. Prevalence estimates and sensitivity analyses**

| **Definition** | **Korea n/elevated; prevalence (95% CI)** | **US n/elevated; prevalence (95% CI)** | **US−Korea difference, pp (95% CI)** |
| --- | --- | --- | --- |
| **Panel A. Primary and age-sex-standardized prevalence** | | | |
| Primary | 5,785/724 12.2% (11.2–13.2) | 1,056/394 35.7% (31.7–39.8) | 23.5 (19.4–27.7) |
| Age-sex standardized | 5,785/724 12.6% (11.6–13.7) | 1,056/394 36.0% (32.1–40.0) | 23.4 (19.4–27.5) |
| **Panel B. Sensitivity analyses** | | | |
| Survey-provided LDL-C <100 mg/dL | 4,751/607 12.5% (11.4–13.6) | 1,083/410 36.2% (32.1–40.3) | 23.7 (19.5–28.0) |
| Sampson LDL-C <70 mg/dL | 2,077/239 11.4% (9.8–13.1) | 340/130 35.9% (29.5–42.4) | 24.5 (17.8–31.1) |
| Non-HDL-C <130 mg/dL | 6,765/823 11.9% (11.0–12.8) | 1,315/484 34.2% (29.9–38.6) | 22.4 (17.9–26.8) |
| hsCRP ≥2 mg/L; no upper cutoff | 5,899/838 13.9% (12.8–14.9) | 1,143/481 41.0% (36.6–45.4) | 27.1 (22.6–31.6) |
| Korea 2022–2023 vs NHANES cycle | 3,732/458 11.8% (10.6–12.9) | 1,056/394 35.7% (31.7–39.8) | 24.0 (19.8–28.2) |
| Treatment proxy absent | 3,348/478 13.6% (12.3–14.9) | 535/204 33.7% (27.6–39.8) | 20.1 (13.9–26.3) |

Counts are unweighted; prevalence and differences are survey weighted. Standardized estimates use the primary samples and the common age-sex reference distribution specified in the main text. Elevated hsCRP is 2–10 mg/L with hsCRP ≤10 mg/L, except the no-upper-cutoff row, where it is hsCRP ≥2 mg/L. Lipid sensitivities retain shared fasting and valid-lipid criteria. pp, percentage points; CI, confidence interval.

**Table S5. Regression estimates and weighting sensitivities**

| **Model / adjustment** | **Common sample PR (95% CI)** | **Model-specific N/elevated** | **Model-specific PR (95% CI)** |
| --- | --- | --- | --- |
| M0 Unadjusted | 2.89 (2.51–3.32) | 6,841/1,118 | 2.92 (2.55–3.36) |
| M1 Age + sex | 2.86 (2.48–3.30) | 6,841/1,118 | 2.92 (2.54–3.35) |
| M2-WC Age + sex + waist circumference | 1.66 (1.40–1.98) | 6,599/1,063 | 1.74 (1.50–2.03) |
| M2-BMI Age + sex + BMI | 1.92 (1.62–2.26) | 6,694/1,088 | 2.03 (1.72–2.39) |
| M2-WC+BMI Age + sex + waist circumference + BMI | 1.68 (1.41–2.01) | 6,514/1,051 | 1.74 (1.50–2.03) |
| M3 Waist model + cardiometabolic covariates | 1.72 (1.43–2.07) | 6,521/1,042 | 1.79 (1.52–2.10) |
| M4 Cardiometabolic model + lifestyle/treatment indicators | 1.74 (1.44–2.10) | 6,314/984 | 1.75 (1.45–2.11) |
| Age + sex, equal total survey weights | — | 6,841/1,118 | 2.96 (2.59–3.39) |

Common samples: N=6,235; elevated hsCRP n=972 in every model. PRs use Korea as reference and design-based CIs. M2-WC, M2-BMI and M2-WC+BMI are alternative adiposity specifications; M3 extends the waist model and M4 extends M3. Covariates are defined in the main text. Common and model-specific estimates use within-survey mean-1 weights. Equal-total weighting is compared with the same model-specific age-sex sample (N=6,841; PR 2.92), not the common-sample PR 2.86.

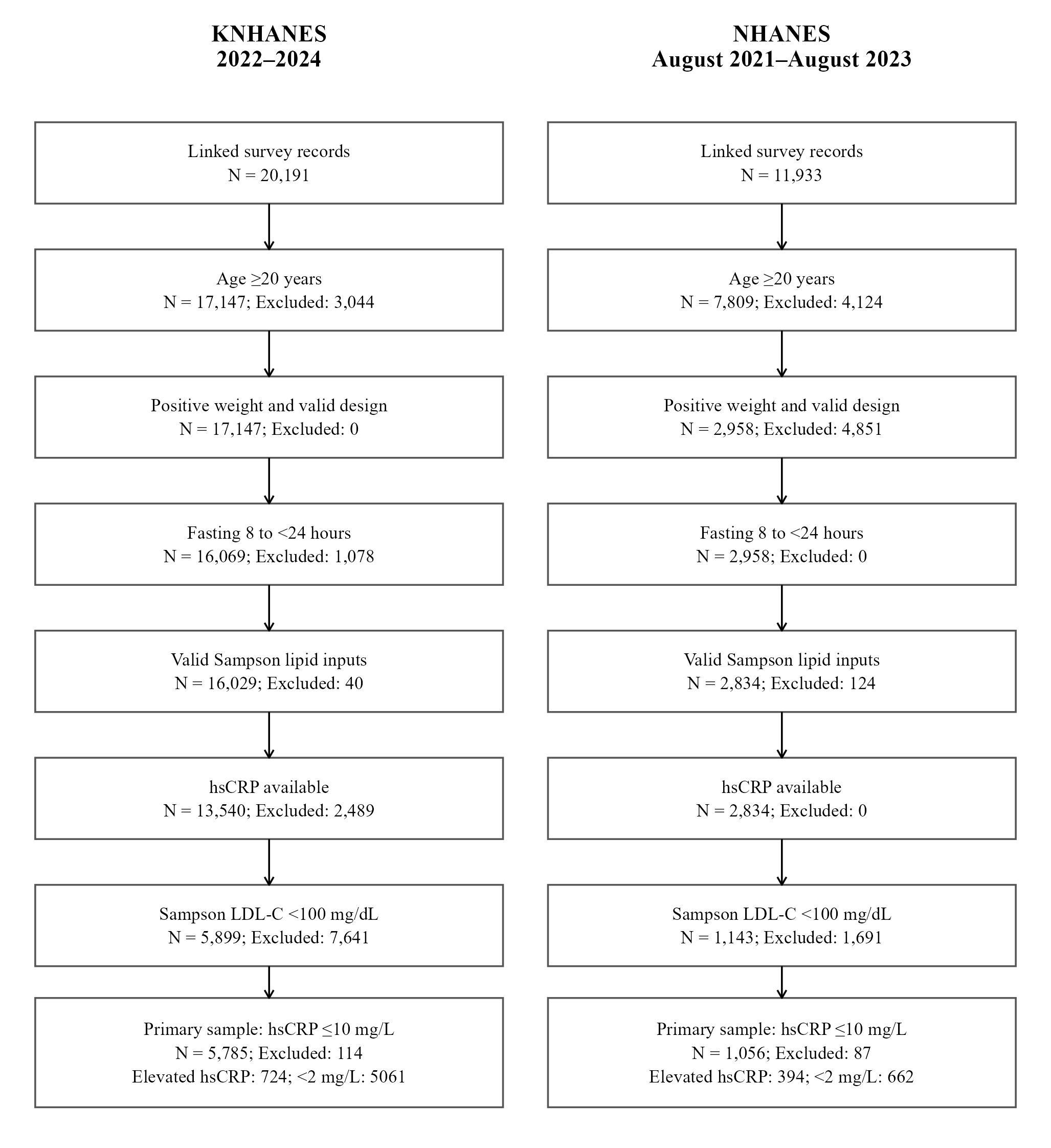

**Figure S1. Derivation of the analytic samples** Counts are unweighted and exclusions are sequential. Common complete cases were constructed separately for regression models.

**Supplementary method references**

S1. CDC. NHANES August 2021–August 2023: TRIGLY_L documentation. https://wwwn.cdc.gov/Nchs/Data/Nhanes/Public/2021/DataFiles/TRIGLY_L.htm (accessed October 7, 2026).

S2. CDC. NHANES August 2021–August 2023: HSCRP_L documentation. https://wwwn.cdc.gov/Nchs/Data/Nhanes/Public/2021/DataFiles/HSCRP_L.htm (accessed October 7, 2026).

S3. Moon S, Kim YJ, Yu JM, Kang JG, Chung HS. Z-score of the log-transformed A Body Shape Index predicts low muscle mass in population with abdominal obesity: the U.S. and Korea National Health and Nutrition Examination Survey. PLoS One. 2020;15:e0242557. doi:10.1371/journal.pone.0242557.

S4. Inker LA, Eneanya ND, Coresh J, Tighiouart H, Wang D, Sang Y, et al. New creatinine- and cystatin C–based equations to estimate GFR without race. N Engl J Med. 2021;385:1737–1749. doi:10.1056/NEJMoa2102953.
